# Cortical and Cortico-Muscular Beta-Gamma Phase Amplitude Coupling during Different Locomotion Status and the Effects of Levodopa in Parkinson’s Disease

**DOI:** 10.1101/2025.04.24.25326354

**Authors:** Haifeng Zhao, Shenglin Hao, Peiran Zhang, Shenghong He, Laura Wehmeyer, Ziyi Feng, Lu Xu, Shikun Zhan, Wei Liu, Xiaoxiao Zhang, Marie-Laure Welter, Dianyou Li, Bomin Sun, Yong Lu, Huiling Tan, Chunyan Cao

## Abstract

**Objectives:** Parkinson’s disease (PD) is a neurodegenerative disorder that can cause motor impairments and gait problems. Non-invasive electrophysiological biomarkers like phase amplitude coupling (PAC) hold promise for understanding and managing PD. This study aims to investigate the modulations of levodopa and locomotion in cortical and cortico-muscular beta-gamma PAC by monitoring central EEG (cEEG) and gastrocnemius EMG (gEMG) and discuss their potential utility as biomarkers for monitoring motor impairments.

**Methods:** The cEEGs and gEMGs were recorded in 30 PD patients during sitting, standing, and free walking under OFF and ON dopaminergic medication. Spectral features, cEEG and cEEG-gEMG PAC were analyzed to assess the effects of levodopa, locomotion, and their correlations with PD symptoms.

**Results:** Longer gait cycle intervals and shorter step lengths were observed OFF levodopa, correlating with UPDRS-III. The cEEG beta-gamma PAC during sitting and standing, and cEEG-gEMG beta-gamma PAC during walking, positively correlated with UPDRS-III OFF levodopa. The cEEG alpha/low beta-gamma and cEEG-gEMG low beta-gamma PAC increased during levodopa withdrawal while walking, with the latter correlating with reduced step length. Step event-related PAC analysis unveiled the enhancement of alpha/beta cEEG-gamma gEMG PAC around heel strikes in ON levodopa compared to OFF.

**Conclusions:** Elevated cEEG and cEEG-gEMG beta-gamma PAC in various locomotion are associated with Parkinsonian motor impairments, whereas cEEG-gEMG low beta-gamma PAC is linked specifically to gait dysfunction. Levodopa administration enhances the dynamic fluctuations of alpha/beta oscillations during free walking. These findings demonstrated that the cortical and cortico-muscular couplings are significantly impacted by both levodopa and locomotion status, hinting the context-dependent nature of PAC and its potential utility in the development of gait phase-locked aDBS strategies for PD patients.

## Graphical Abstract

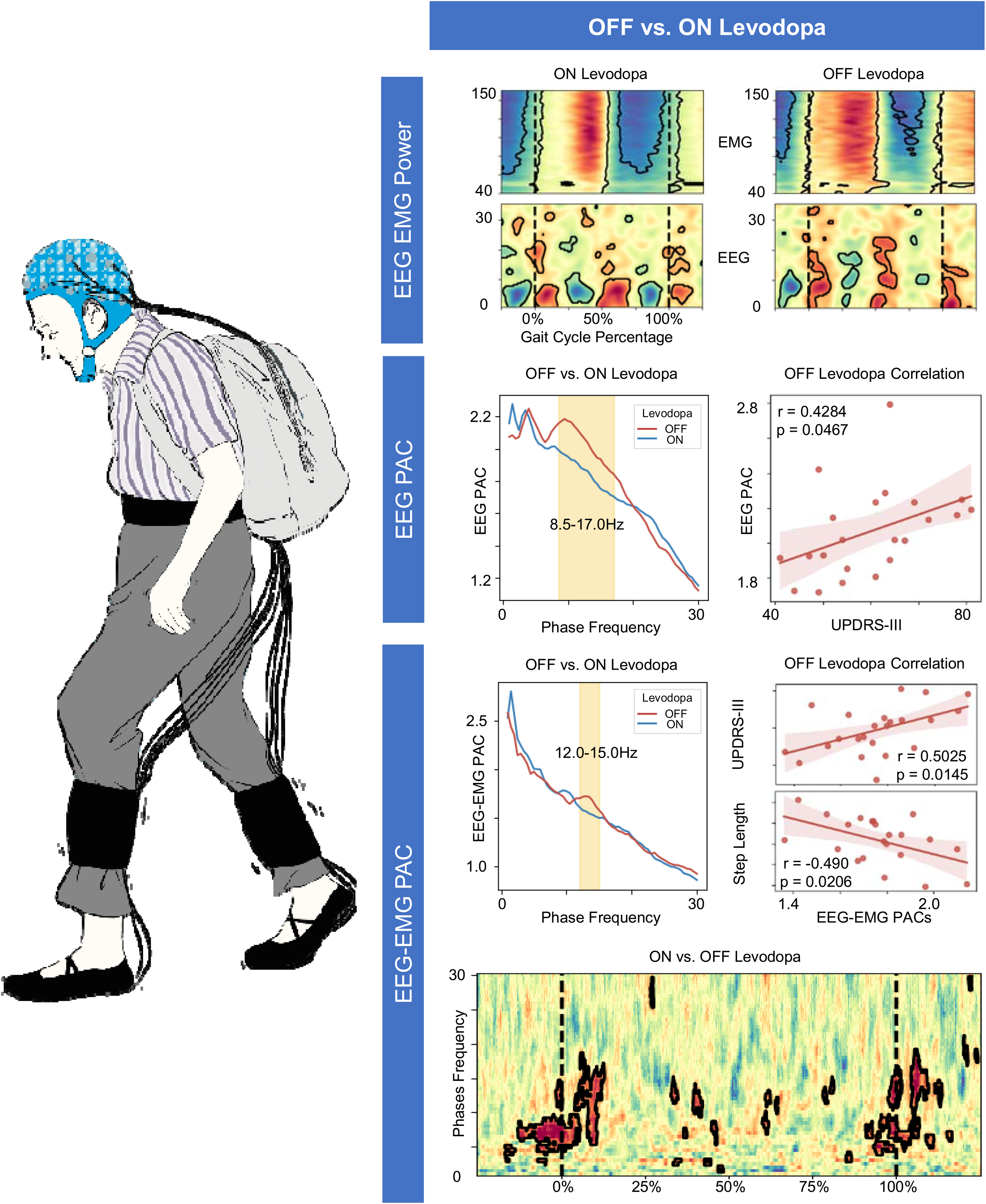

## 1. Introduction

Parkinson’s disease (PD) impairs motor function due to dopaminergic neuron loss in the substantia nigra, leading to symptoms like tremor, bradykinesia, rigidity, and postural instability. Whilst clinical evaluation by medical doctors remains the gold standard for PD diagnosis and assessment^1,2^, it is highly dependent on clinician’s experience and lacks continuous monitoring of symptom fluctuations throughout the day^3^. Electrophysiological biomarkers, such as beta and gamma oscillations in subcortical local field potentials (LFPs), EEG or magnetoencephalography (MEG) measurements, and EMG, offer objective insights into PD-related neural disruptions and hold promise for diagnosis and assessment^4^.

Elevated beta power in basal ganglia, such as subthalamic nucleus (STN) and globus pallidus interna (GPi) has been well established as a hallmark for PD^5,6^. Abnormal beta bursts are associated with motor symptoms like bradykinesia and rigidity^7^, reflecting pathological beta hyper-synchronization that disrupts normal motor function^5,8^. Meanwhile, fine-tuned gamma activity becomes more prominent with dopamine therapy and can be entrained by deep brain stimulation (DBS)^9,10^. These insights have guided therapeutic strategies like DBS, which aim to reduce beta and enhance gamma activity in the basal ganglia^5,11–13^. However, beta/gamma power changes in the cortex, particularly when measured non-invasively using EEG or MEG, remain inconsistent^14–18^.

Meanwhile, phase amplitude coupling (PAC), which describes the modulation of higher-frequency oscillations by the phase of lower-frequency rhythms, holds promise for understanding cross-frequency interactions in the brain. Previous studies have shown that PAC in the STN and GPi, involving motor-relevant frequencies, such as beta, coupled with high-frequency gamma (40-200Hz) oscillations, reflects the pathological synchronization seen in PD^19–23^. Several studies have suggested enhanced beta-gamma PAC in the primary and sensorimotor cortex as a biomarker for PD as they correlated with PD motor symptoms^24–26^, and reduced with dopamine medication^27^ and therapeutic deep brain stimulation^28^. Increased cortical beta-gamma PAC has also been implicated in freezing of gait (FOG), with one study using ECoG reported increased beta-gamma PAC in the motor cortex during the trials with freezing of gait (FOG)^29^. Another study recorded scalp EEG in the preparation for and during gait showing that beta-gamma PAC in the sensorimotor area during preparation for walking differs depending on the emergence of FOG. As gait symptoms worsened, beta-gamma PAC in the sensorimotor area during walking gradually increased^30^. Therefore, it has been suggested that cortical PAC may serve as a biomarker for FOG in PD for the development of new strategies to prevent falls in the future.

Despite the recent interest in cortical beta-gamma PAC as a biomarker for PD symptoms and FOG, it is still not clear how they are modulated by different locomotion states and how the modulation is changed by dopaminergic medication. In addition, the specific mechanisms by which cortical activity couples to muscle output remain insufficiently understood. Limited studies^31–34^ investigated cross-region coherence among cortex, basal ganglia and muscles, while cross-frequency coupling between cortex and muscle remains unexplored. For example, reductions in beta band cortico-muscular coherence (CMC) in PD have been reported ^32,33^, whereas increases in gamma band CMC have been observed during muscle contractions^35^ and dynamic force controls^36^. However, cross-frequency cortico-muscular coupling in different locomotion contexts has not been studied, and may offer new insight on how cortical activity couples to muscle output during standing and walking.

Our study addresses these knowledge gaps by employing simultaneous non-invasive EEG and EMG recordings across multiple locomotion states (sitting, standing and free walking) in PD patients with medication both ON and OFF, with footpad sensors tracking different events (such as heel-strike and toe-off) within each step-cycle during free walking. This allows us to investigate the role of beta-gamma PAC in the central EEG (cEEG) across different motor states and explore how step-related events, such as heel strikes, modulate PAC within each step cycle. We also aim to assess correlations between PAC and clinical measures of motor disability and gait instability, and examine how levodopa administration modulate cortical PAC as well as cortico-muscular couplings.

## 2. Methods

### 2.1 Participants and Ethical Approval

The study was approved by the Ethics Committee of Ruijin Hospital, Shanghai Jiao Tong University School of Medicine. Thirty participants with PD (18 males), aged 52 to 76 years (mean age 63.97, SD 6.97), took part in the study. The participants were assessed under both OFF and ON dopaminergic treatments, as indicated in Table 1. The Movement Disorder Society revised Unified Parkinson’s Disease Rating Scale part III (UPDRS-III) varied across participants from 26 to 101 in the OFF state and from 9 to 68 in the ON state, with a mean improvement of 41.34% following dopaminergic treatment (Table 1). The UPDRS-III assessments were performed by professional clinicians prior to the study according to MDS-UPDRS guidelines set by the International Parkinson and Movement Disorder Society. The scale was organized into specific subsections for detailed analysis, such as rigidity (3.2-3.3), akinesia (3.4-3.14), tremor (3.15-3.18) and gait problems (3.10-3.14).

### 2.2 Task Paradigm

Figure 1A illustrates the three locomotion states (sitting, standing, and walking) during which EEG and EMG data were recorded under both OFF and ON levodopa conditions. The OFF levodopa state was defined as at least 12 hours of medication withdrawal, while the ON levodopa state was assessed 1-2 hours after levodopa intake of a challenge dose (150% see Table 1 for details). Participants sat quietly for over one minute, and then stood for over one minute. For the walking condition, they walked back and forth over a distance of five meters for 5 minutes. One-minute continuous data segments were extracted from each locomotion state for overall power and PAC analysis. In addition, 20 clean straight-walking steps per side were segmented into epochs using foot-pressure triggers. All participants completed the sitting and walking tasks in the ON levodopa state, while only 24 completed the standing task. In the OFF levodopa condition, 26 participants completed the sitting and walking tasks, and 21 completed the standing task.

**Figure 1.**
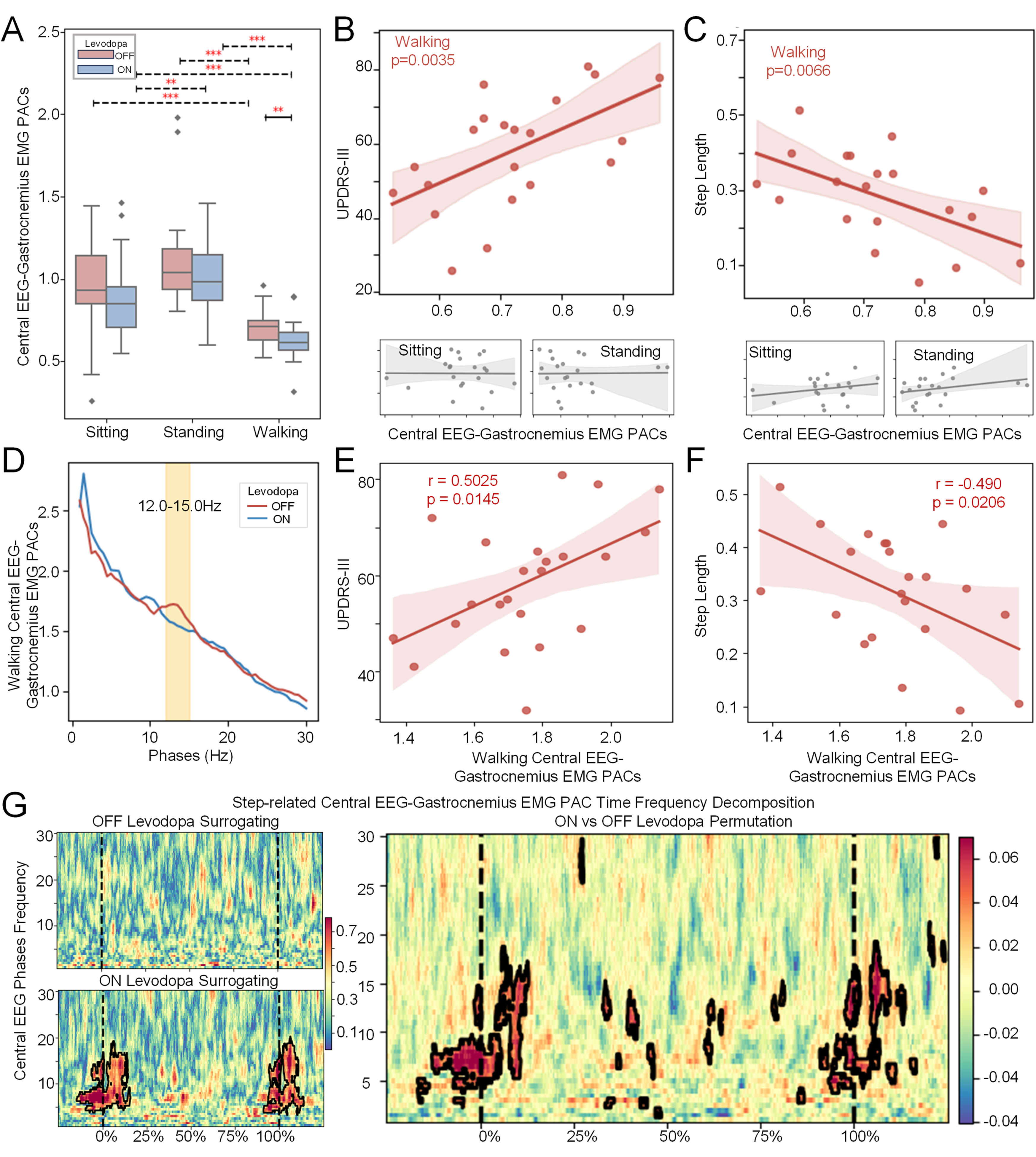
**(A) Task Paradigm:** Each participant was recorded during sitting, standing, and walking, and this was repeated under both OFF and ON levodopa conditions. One-minute segments and the first 20 steps of walking were analyzed to compare dynamic gait patterns and levodopa effects. **(B) Device Setup:** The system monitored neuro-electrophysiological signals during movement. An EEG cap captured brain activity, and EMG sensors placed on the gastrocnemius muscle recorded lower limb muscle activity. Signals were synchronized with foot pressure triggers and processed by an Arduino microcontroller, ensuring accurate step recording. **(C) Step Identification:** Foot pressure sensors placed at the toe and heel were utilized during walking. Heatmap plot indicates that heel strikes were more consistent, establishing heel pressure as a reliable measure for step analysis. The bottom figure displays the pressure signal (blue) and the associated phase (orange) plot. Red dotted lines indicate thresholds for determining the events of heel strike (3V) and release (4.5V). **(D) Channel Positions:** The EEG setup followed the 10-20 system, with additional electrodes at C1 and C2. The analysis focused on channels C1, C2, and Cz, located over the central cortex, which are crucial for processing sensory and motor signals from the lower limbs.

### 2.3 Data Recording

Figure 1B illustrates the setup integrating EEG, EMG, and digital triggers in real time. EEG was recorded using a 10-20 system cap with an embedded reference electrode (FCz), and additional electrodes at C1 and C2, totaling 23 channels. For analysis, channels C1, C2, and Cz, located near the primary motor cortex (M1), were selected due to their established relevance to lower limb motor functions^37–40^. The remaining channels were used to ensure signal quality, support noise removal, and allow for channel interpolation when needed. EMG signals were recorded from the gastrocnemius muscle, which is crucial for maintaining balance during the heel strike^41^ and plays a vital role during the push-off phase. These neuro-electrophysiological measurements are referred to as cEEG (central EEG: C1, C2, Cz) and gEMG (gastrocnemius EMG) throughout the manuscript. Both EEG and EMG were sampled at 512Hz using the Compumedics Neuroscan Grael EEG 2 system. Foot pressure data was processed by an Arduino microcontroller at 16MHz. Figure 1C shows an example where a 3V threshold was conclusively identified as the heel strike (rather than toe strike), ensuring robust step detection.

### 2.4 Data Processing

Initial analyses were conducted on one-minute continuous data for each locomotion state (sitting, standing, and walking). Walking data was then further segmented into individual steps using footpad pressure sensor triggers. The gait cycle interval was defined as the time between consecutive heel strikes from the same foot. Since step length could not be measured directly by the sensors, video recordings were used to count the number of steps between two fixed five-meter landmarks, with the mean step length calculated by dividing the distance by the number of steps. All EEG data were filtered, average-referenced, and cleaned using independent component analysis (ICA) to remove non-brain artifacts components and retain ‘brain’ components as labeled by ICLabel^42^. To reduce neck-related and movement artifacts on the for analyses selected central lobes channels, additional artifact rejection criteria, including peak-to-peak thresholds and mean/std outlier removal, were applied to exclude noisy epochs. Subsequently, Time-frequency representations (TFRs) were visually inspected to ensure that no cross-frequency artifacts were included in the analysis. All further analysis were performed on channel level and then averaged. All event-related analyses were time-locked to heel strikes, using EMG from the ipsilateral muscles and EEG from the contralateral hemisphere. The processing was done by using MNE-Python^43^. Detailed descriptions of the preprocessing methods are provided in the supplementary materials.

### 2.5 Phase Amplitude Coupling

To investigate PAC, we employed the Gaussian Copula PAC (GC-PAC) method from Tensorpac^44^, which is well-suited for analysing short-duration data between different signal sources:

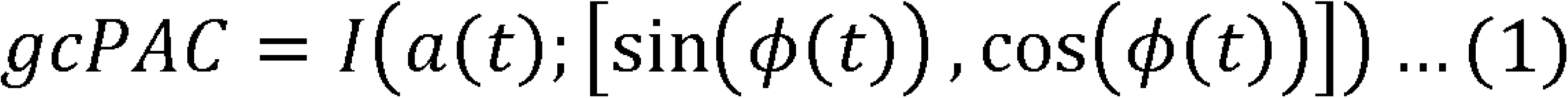

The method first applies a copula normalization to transform the phase and amplitude data into a standard normal distribution. This step mitigates variations in amplitude scaling and signal-to-noise ratios across modalities, allowing for a robust, bias-corrected Gaussian estimation of mutual information. The step-related PAC was calculated using the Event-Related PAC (ERPAC) measure, which employs a circular-linear correlation approach. This method evaluates the Pearson correlation across trials between the amplitude and the sine and cosine of the phase:

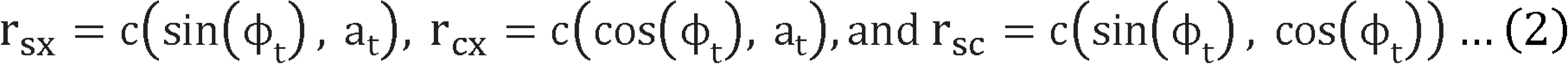

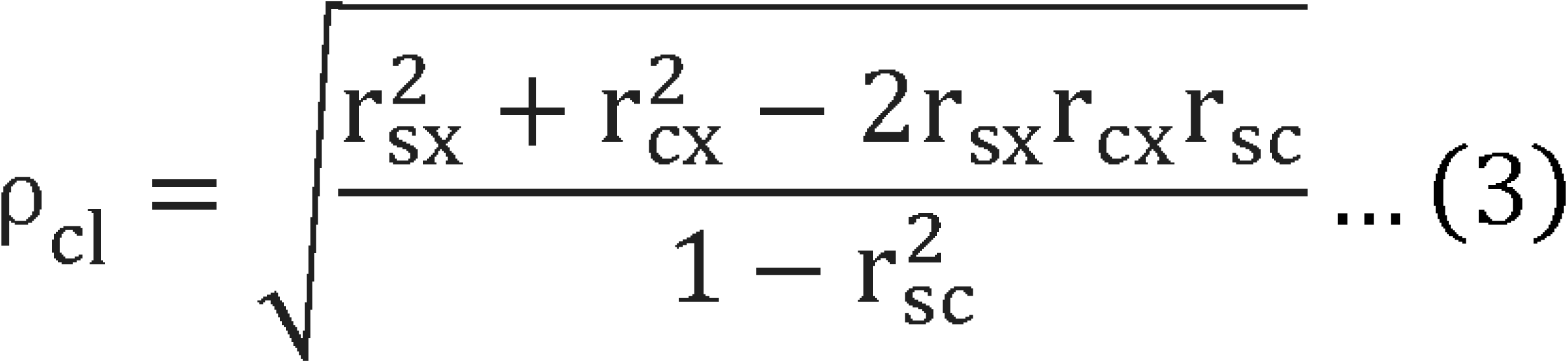

### 2.6 Statistical Analysis

We employed two complementary approaches: an evidence-based method that replicated baseline results using established bandwidths (e.g., alpha: 8–12Hz, beta: 12–30Hz, and gamma: 40–200Hz) from previous studies^29,45^, and a data-driven method using permutation and surrogate testing to identify frequency bands sensitive to specific levodopa and locomotion conditions. A two-way ANOVA was conducted to assess the main and interaction effects between levodopa condition (OFF vs ON) and locomotion state (sitting, standing, walking) on the powers and PAC. For post-hoc comparisons, the non-parametric test Mann-Whitney U was employed. A cluster-based permutation test with 2000 iterations was employed to identify statistically significant differences in powers/PAC. EEG and EMG time-frequency decompositions were permuted between walking steps and random, same-length segments from sitting. The original data was permuted at the step segment level between OFF and ON conditions, and then PAC was re-calculated to generate a null distribution. Significant step-related PAC clusters in each levodopa condition were identified by comparing the target PAC with its phase-signal surrogates. We first used a Generalized Linear Model (GLM) to examine the association between measured gait symptoms and UPDRS-III attributes. Subsequently, the GLM was employed to identify which PAC measure—derived from different locomotion conditions—best predicts motor symptoms, including UPDRS-III scores, step length, and gait cycle intervals. Pearson correlation assessed linear relationships between variables. Z-score outlier removal with a threshold of three was used to ensure data quality by eliminating data points. The False Discovery Rate (FDR) was controlled for multiple comparisons using the Benjamini-Hochberg. The original results were reported in the figures and results with FDR-corrected p-values specified in annotations.

## 3. Results

### 3.1 Behaviors Measurements

The Mann-Whitney U test showed a significantly longer gait cycle interval (*U*=491.0, *p*=0.0285, Figure 2A) a\nd shorter step length (*U*=182.0, *p*=0.0028) in the OFF compared to ON levodopa state (Figure 2C). Pearson correlation analyses revealed that both gait cycle intervals (*r*=0.4243, *p*=0.0308, Figure 2B) and step length (*r*=-0.5797, *p*=0.0024, Figure 2D) correlated with UPDRS-III scores in the OFF state, indicating significant influence of PD symptoms on these walking patterns. GLM regression further showed that the gait cycle interval increase might be ascribed to rigidity (β=0.0140, *p*=0.0405, FDR-adjusted *p*=0.1620), although this did not survive correction, whereas shorter step length remained significantly linked to higher UPDRS-III subsection gait scores (β=1.6998, *p*=0.0018, FDR-adjusted *p*=0.0073). Levodopa treatment significantly improved motor performance, with an average 41.34% reduction in UPDRS-III score. The improvement was observed across various motor symptoms, with reductions of 38.24%, 41.50%, 44.50%, and 53.27% in rigidity, akinesia, tremor, and gait, respectively (*p*<0.01).

**Figure 2.**
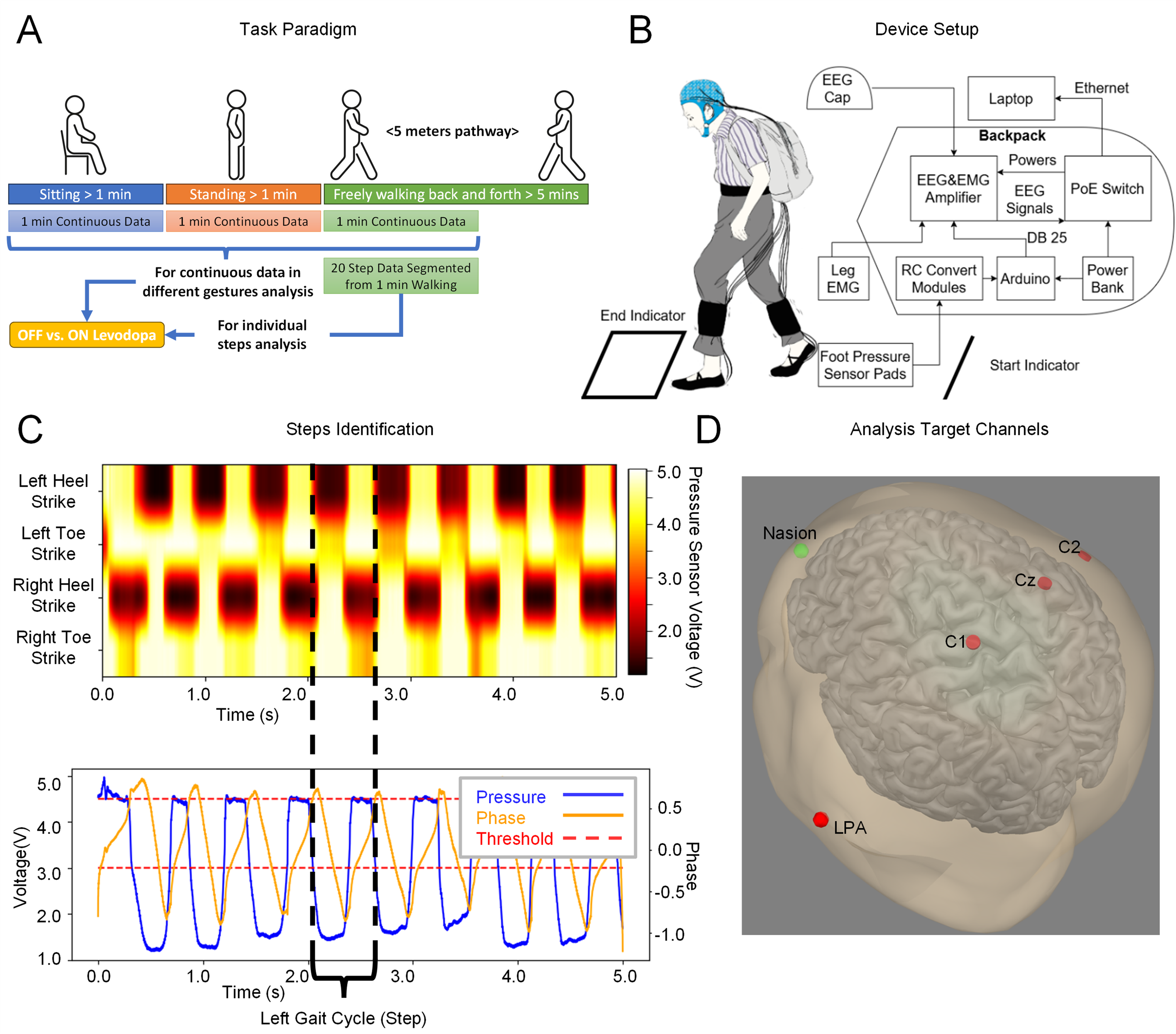
Statistics of Gait Cycle Intervals and Step Length. (A) The gait cycle intervals in the ON levodopa state are significantly shorter than those in the OFF levodopa state. (B) The gait cycle intervals in the OFF levodopa state show a significant correlation with UPDRS-III scores. (C) The step lengths in the OFF levodopa state are significantly shorter than those in the ON levodopa state. (D) The step lengths in the OFF levodopa state are significantly correlated with UPDRS-III scores.

### 3.2 Central EEG and Gastrocnemius EMG Powers

A two-way ANOVA indicated significant main effects of levodopa (*F*=7.5470, *p*=0.0070) and locomotion (*F*=220.8337, *p*<0.0001) on gEMG gamma band (40-200Hz) as well as a significant interaction effect between levodopa and locomotion (*F*=5.0968, *p*=0.0075). Post-hoc tests confirmed a substantial decrease in gEMG power during the ON compared to OFF levodopa condition while sitting (*U*=393.0, *p*=0.0047, FDR-adjusted *p*=0.0061), whereas no significant differences were observed during standing (*U*=140.0, *p*=0.4408) or walking (*U*=248.0, *p*=0.7332). Movement significantly increased gEMG power from sitting to standing and then walking in both OFF and ON Levodopa conditions (*p*<0.0001, FDR-adjusted *p*<0.0001). Conversely, a two-way ANOVA revealed a significant effect of locomotion on cEEG alpha (*F*=3.1583, *p*=0.0459) and broad gamma band power (*F*=7.2906, *p*=0.0010). No significant levodopa effects were observed in any specific band. Alpha power was reduced during walking compared with sitting, especially when ON medication (*U*=439.0, *p*=0.0127, FDR-adjusted *p*=0.1145, Figure 3A). In contrast, gamma band power increased during walking compared to sitting in both ON (*U*=186.0, *p*=0.0034, FDR-adjusted *p*=0.0308, Figure 3A) and OFF levodopa conditions (*U*=160.0, *p*=0.0228, FDR-adjusted *p*=0.1028, Figure 3A). The PSD analysis of the cEEG revealed a decrease within the alpha and beta bands as participants transitioned from sitting to standing to walking, but permutation testing revealed no frequency-specific differences reaching statistical significance (Figure 3B). In addition, we investigated how the cEEG and gEMG activities were modulated by walking gait cycles. Significant modulations were observed in alpha and beta band power in the cEEG within a single gait cycle during the ON levodopa condition, with both alpha and beta activity decreasing before the contralateral heel strike and increasing afterward. In the OFF condition, this modulation was more diffusely distributed across both time and frequency domains. (Figure 3C).

**Figure 3.**
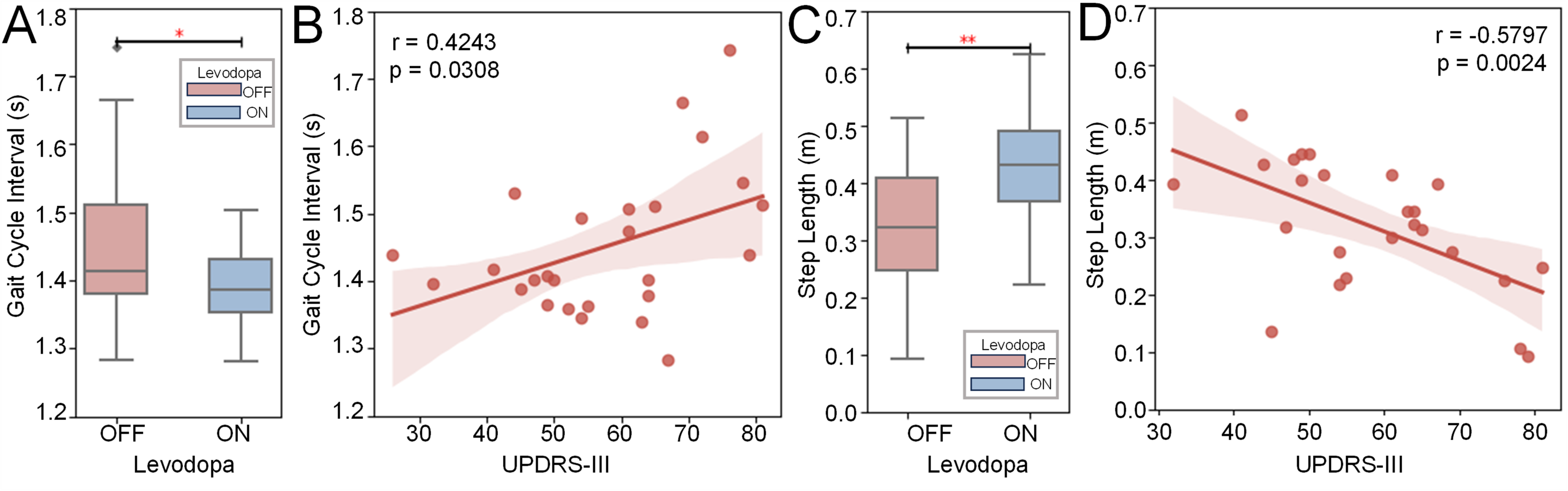
EMG and EEG Power Analysis. (A) For the averaged EEG power analysis across C1, C2, and Cz channels, only locomotion but not medication leads to significant changes in power. Alpha band power significantly decreases during walking compared to sitting at rest in the ON levodopa condition. Conversely, gamma band power significantly increases during walking in both the OFF and ON levodopa states. All values are processed with log10 + 5 for improved presentation. (B) Power Spectrum Density (PSD) of Central Lobes EEG: The PSD in alpha and beta bands decreases from sitting to standing to walking but no significant differences were observed amongst various locomotion or between OFF and ON levodopa status. (C) Gait cycle power spectra aligned to heel strikes as starter (time 0) showed a prominent event-related modulation especially in the alpha and beta frequency band during ON levodopa. EEG powers were averaged over contralateral side of heel strike (C2, Cz for the left heel strike, C1, Cz for the right heel strike), then further averaged between two side of the body. EMG was recorded from the ipsilateral gastrocnemius of heel strikes. The gait cycle percentage was demonstrated only on lateral side for better presentation, with blue indicating right foot and green the left. The start and end time point corresponds to consecutive heel strikes, and the power spectrum was z-score normalized before plotting.

### 3.3 Central EEG PAC

We observed an increase in cEEG beta-broad gamma PAC in the OFF levodopa condition compared to the ON levodopa condition, particularly during standing (Figure 4A). Specifically, phase modulation was most prominent around 10Hz. However, to maintain consistency with previous studies, we initially adopted a hypothesis-driven approach, focusing on a beta frequency range (12-30Hz) for phase and broad gamma range (40-200Hz) for amplitude. A two-way ANOVA on data within these frequency bands confirmed a significant main effect of levodopa (*F*=4.7642, *p*=0.0306) and interaction effect between levodopa and locomotion (*F*=4.2484, *p*=0.0161), whereas the main effect of locomotion alone was not significant (*F*=2.6811, *p*=0.0718). Specifically, levodopa significantly reduced beta-gamma PAC, with the effect most pronounced during standing, although this reduction did not survive FDR correction (*U*=339.0, *p*=0.0227, FDR-adjusted *p*=0.0680, Figure 4B). In the OFF levodopa state, beta-gamma PAC during standing increased significantly compared to sitting (*U*=148.0, *p*=0.0119), although the FDR-adjusted p-value (0.0537) did not reach significance. Only when ON levodopa did beta-gamma PAC during walking decrease significantly relative to sitting (*U*=625.0, *p*=0.0041, FDR-adjusted *p*=0.0366). We also found UPDRS-III scores increased significantly with higher PAC during sitting and standing but not walking in cEEG in OFF condition (β=43.9625, *p*=0.0302, FDR-adjusted *p*=0.0453; β=19.0411, *p*=0.0203, FDR-adjusted *p*=0.0453; β=-1.1020, *p*=0.8914, Figure 4C). To explore the frequency with the most significant phase modulation, PAC was averaged across the broad gamma range (40-200Hz). This analysis identified a significant difference in the phase modulation frequency band from 8.5Hz to 17.0Hz (*p*<0.05, Figure 4D), corresponding to the alpha/low-beta frequency band. This PAC was subsequently averaged across phase frequencies for post-hoc statistical test which revealed a significant increase in PAC under OFF levodopa conditions compared to ON during walking (*U*=417.0, *p*=0.0340). The Pearson correlation analysis showed a significant correlation between alpha/low beta (8.5-17.0 Hz) to broad gamma (40-200Hz) PAC and UPDRS-III (*r*=0.4284, *p*=0.0487, Figure 4E).

**Figure 4.**
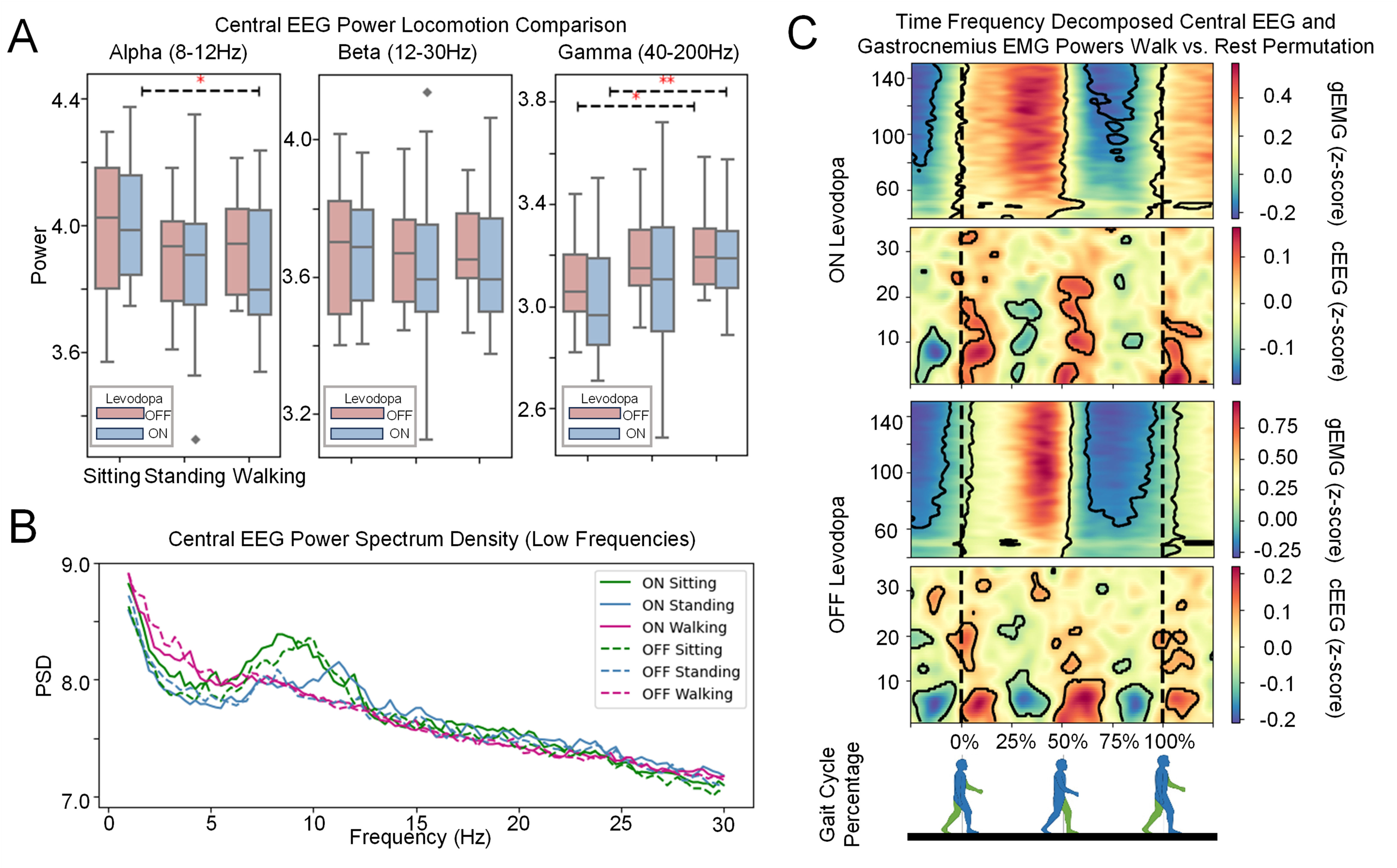
PAC results for cEEG. The PAC values were processed with log10 + 5 for improved presentation. (A) The OFF vs ON PAC during various locomotion illustrates a high contour around alpha/beta phase and gamma amplitude(B) A two-way ANOVA reveals a significant effect of levodopa and an interaction between levodopa and movement state on cortical beta (12-30Hz) to gamma (40-200Hz) PAC. Post-hoc test further showed significant differences between OFF and ON states when standing. (C) A GLM-based multiple regression model demonstrates that OFF levodopa sitting and standing PAC correlated with UPDRS-III scores in the OFF levodopa state. (D) The frequency decomposed walking cortical PAC were further averaged across 40-150Hz in amplitudes dimension to explore the significantly different phase frequencies. The permutation test reveals that during walking cortical PAC at phase frequencies of 8.5-17.0Hz is significantly higher in the OFF levodopa state compared to the ON levodopa state. (E) OFF levodopa averaged alpha/low beta to broadband gamma PACs during walking were significantly correlated with UPDRS-III overall scores (*p* = 0.0467).

### 3.4 Central EEG-Gastrocnemius EMG PAC

Both levodopa and locomotion had main effect on cEEG-gEMG beta-gamma PAC (*F*=4.9622, *p*=0.0274; *F*=45.2271, *p*<0.0001). However, no interactive effect between levodopa and locomotion was observed (*p*=0.8929, Figure 5A). Post-hoc test revealed the significant reduction impact of levodopa on cEEG-gEMG PAC only during walking (*U*=532.0, *p*=0.0092, FDR-adjusted *p*=0.0138), with no comparable reduction observed during sitting or standing (*p*=0.1553; *p*=0.3451). The cEEG-gEMG beta-gamma PAC decreased significantly during walking compared to both sitting (*U*=535.0, *p*=0.0003, FDR-adjusted *p*=0.0007, OFF; *U*=737.0, *p*<0.0001, FDR-adjusted *p*<0.0001, ON) and standing (*U*=530.0, *p*<0.0001, FDR-adjusted *p*<0.0001, OFF; *U*=665.0, *p*<0.0001, FDR-adjusted *p*<0.0001, ON), independently of levodopa state. In the OFF levodopa state, GLM demonstrated that increased cEEG-gEMG beta-gamma PAC during walking was significantly associated with higher UPDRS-III scores (β=76.0980, *p*=0.0035, FDR-adjusted *p*=0.0106, Figure 5B), whereas no such association was observed during sitting or standing (β=-3.7152, *p*=0.7560; β=5.9677, *p*=0.5615). Moreover, the average cEEG-gEMG PAC during walking negatively correlated with step length in the OFF levodopa state, where increased cEEG-gEMG beta-gamma PAC was associated with shorter step lengths during walking (β=-0.5576, *p*=0.0066, FDR-adjusted *p*=0.0197, Figure 5C). However, GLM analysis did not reveal any significant associations between step length and cEEG-gEMG beta–gamma PAC during sitting or standing (β=0.1149, *p*=0.2185; β=0.0582, *p*=0.4653, Figure 5C). When examining the decomposed PAC averaged across gEMG amplitude frequencies (40-200Hz), significant modulation phases were confined to the 12.0-15.0Hz range, corresponding to the cEEG low beta frequency (Figure 5D). Further paired analysis confirmed significant differences between OFF and ON levodopa conditions (*U*=478.0, *p*=0.0207) during walking in the cEEG low beta (12.0-15.0Hz) and gEMG broad gamma (140.0Hz-200.0Hz) PAC. Moreover, cEEG-gEMG PAC in the OFF condition significantly correlated with UPDRS-III scores (*r*=0.5025, *p*=0.0145, Figure 5E). This frequency-specified cEEG-gEMG PAC also significantly correlates with step length (*r=*-0.490, *p=*0.0206, Figure 5F). Figure 5G presents step-related PAC during the gait cycle. Surrogate testing revealed significant time-locked contours in the ON-levodopa condition, indicating increased cEEG alpha/beta-gEMG gamma PAC during contralateral heel strike (*p*<0.05). The permutation test confirmed the significance of these increases during ON levodopa compared to OFF condition.

**Figure 5.**
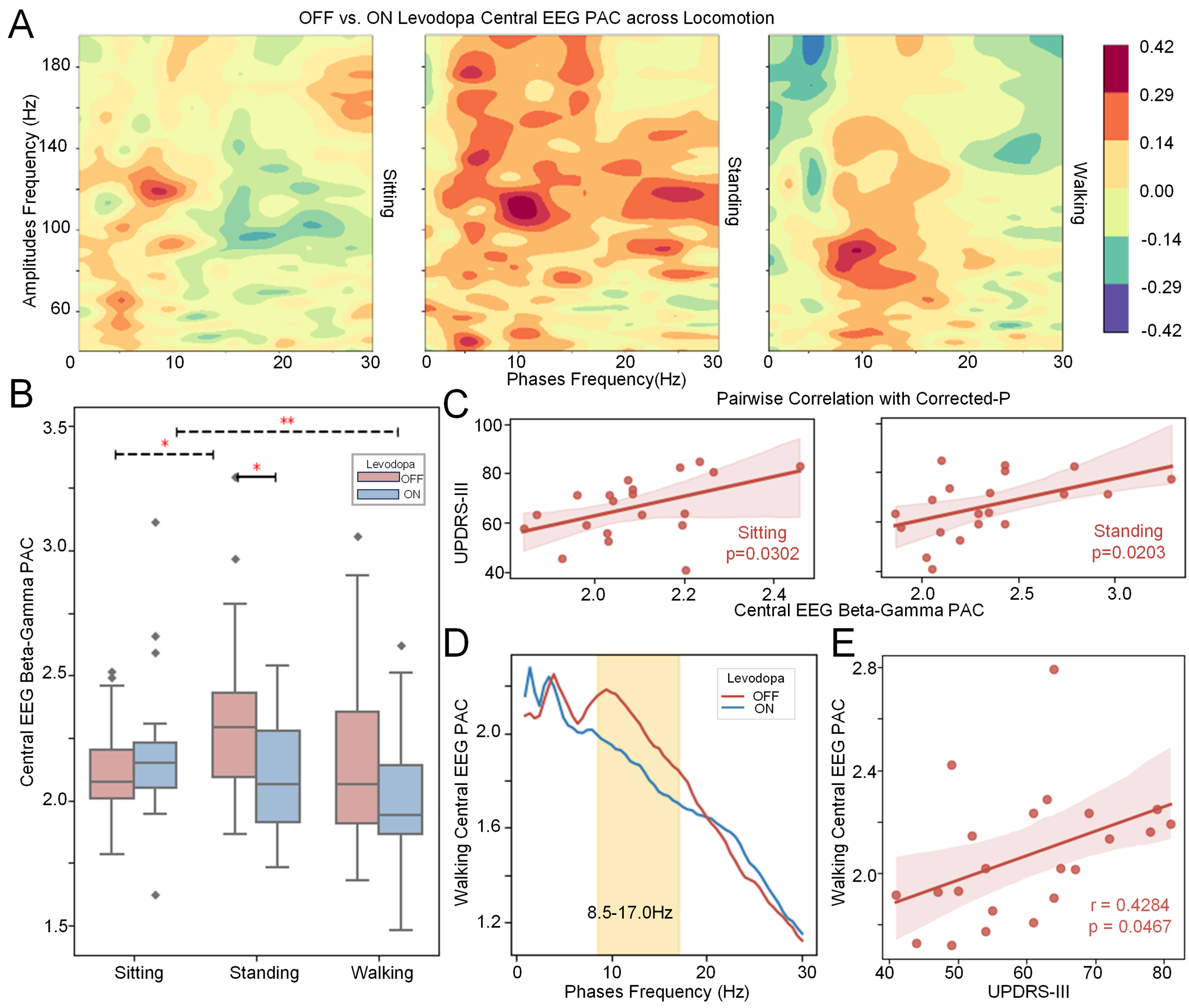
PAC results for cEEG-gEMG. The PAC values are processed with log10 + 5 for better presentation. (A) A comparison of averaged beta (12-30Hz) to broadband gamma (40-200Hz) PACs across levodopa and gesture conditions shows a significant reduction in cortico-muscular PACs during walking in the ON levodopa state compared to the OFF levodopa state, with walking exhibiting lower PACs than sitting in both states. (B) GLM analysis reveals a significant correlation between OFF levodopa walking PACs and UPDRS-III scores but not sitting and standing. (C) GLM analysis also shows the walking step length is specifically correlated with walking cEEG-gEMG PAC during OFF conditions. (D) Analysis of phase frequencies highlights a 12-15Hz low beta band difference between levodopa states in walking cortico-muscular PAC. (E) The data-driven determined phase frequency band walking cEEG-gEMG PAC shows significant correlations to UPDRS-III in the OFF levodopa state(p=0.0145). (F) Frequency-specified walking cEEG-gEMG PACs in the OFF levodopa state significantly correlate with the walking step length (p=0.0206). (G) Step-based PAC analysis shows significant cEEG-gEMG alpha/beta-gamma coupling around heel strikes. The permutation test confirmed these increased modulations are significant in ON than OFF levodopa.

## 4. Discussion

This study introduces a novel, noninvasive system to monitor cortical activities and cortico-muscular interactions in PD patients during sitting, standing and free walking. Results of the study confirmed the pathological significance of cortical beta-gamma PAC, as well as cortical beta-gastrocnemius EMG (gEMG) gamma PAC, in the motor symptoms and gait impairments in PD, with both of these cross-frequency couplings increased in the OFF levodopa condition, correlated with UPDRS scores and reduced step length during free walking. These findings are consistent with previous studies reporting increased cortical beta-gamma PAC in PD, measured via EEG, MEG^25,26,46,47^ or ECoG^29,48^, supporting its potential role as a potential biomarker for motor impairment and FOG. We also revealed dynamic fluctuation of cortical beta-EMG gamma PAC within each step cycle when the patients are ON levodopa with cortico-muscular beta-gamma PAC relatively increased around heel strikes, whereas this cortico-muscular coupling remained high and stable within each step cycle without dynamic fluctuation when OFF medication. These results extend existing literature by offering new insights into how cross-frequency and cross-region coupling are modulated not only by levodopa but also different locomotion states and within each step cycle during free-walking.

### Locomotion-status-dependent cortical PAC reflects levodopa effects and PD’s symptom severity

Elevated cortical alpha/low beta to gamma PAC during sitting has been associated with higher UPDRS-III scores, as shown in prior work^49^. In addition, Yin et. al.’s^29^ observed enhanced low beta-gamma PAC in the motor cortex during standing and at the onset of FOG. This enhancement was attributed to beta hyper-synchrony in the basal ganglia, which likely drives stronger beta-gamma coupling in the motor cortex^50^. Our results also suggest that cEEG beta-gamma PAC is a more reliable predictor for UPDRS scores when quantified during sitting and standing than during walking. This is consistent with previous studies^29,30,51,52^ and suggests that cEEG PAC may serve as a more stable indicator of levodopa responsiveness and symptom severity in static postures. In addition, our data-driven approach highlighted a broader modulatory frequency band, including both alpha and low-beta (8.5-17.0Hz), modulating broad gamma activity (40-200Hz) in the cortex. This PAC is significantly higher when OFF levodopa compared to ON levodopa during walking and correlated with motor symptom severity, reflected as higher UPDRS-III. These findings suggest that the frequency bandwidth of PAC changes associated with symptom severity and levodopa effects may vary depending on the locomotion context.

### Cortical beta-muscular gamma PAC during free walking is modulated by levodopa and its overall elevation indicates severe motor impairment but selective enhancement may play a functional role

Interestingly, cortico-muscular beta-gamma PAC was reduced during walking compared to sitting and standing, and this reduction appeared independent of levodopa condition. In previous studies, cortical beta was found to be “promoter of status quo”^5,53^ whereas gamma rhythms track active muscle contraction^36^. The reduction of cortico-muscular beta-gamma PAC during walking reported here may reflect a functional uncoupling of cortical beta to muscular gamma to permit dynamic motor adjustments. And the loss of dopamine may increase aberrant cortico-muscular beta-gamma coupling during walking. The further GLM confirmed this increase associated with severe motor impairment (higher UPDRS-III) and worse gait (shorter step length). These findings suggest that cortico-muscular beta-gamma PAC during walking appears to be a key marker of both levodopa status and symptoms severity. Furthermore, our data-driven approach narrowed down the phase frequency range to a low beta frequency (12-15Hz) that cortical low-beta and muscular gamma PAC was specifically elevated during walking in the OFF levodopa condition and positively correlated with motor symptom severity, highlighting its role in the pathophysiology of free walking.

In addition, we observed dynamic fluctuation in the cortico-muscular low beta-gamma PAC within each gait cycle only in the ON levodopa condition (Figure 5G), whereas there is lack of such modulation pattern locked to the heel-strikes when OFF medication. This suggests that selective enhancement in cortico-muscular PAC within a gait cycle may play a key functional role under dopaminergic influence. This is consistent with previous findings showing that activities in the STN^54^, PPN^45^, and motor cortex are dynamically modulated within each step cycle. Levodopa suppresses cortico-muscular beta-gamma PAC baseline and restores dynamic cortical beta gating. As a result, phasic beta-gamma PAC peaks reappear at key stage like heel strike, facilitating precise muscle control. PAC enhancements occurring in different temporal contexts may have distinct functional implications. Our results align with previous work suggesting that temporally-targeted interventions, such as alternating DBS^55,56^ or gait phase-locked DBS, may offer improved therapeutic outcomes for gait impairments in PD.

### Limitations

This study has several limitations. First, recordings were relatively short and confined to a laboratory setting. It remains to be tested whether the reported results generalize to everyday life, where motor behavior is more diverse. Second, we used central lobe EEG to noninvasively monitor cortical activity over the motor regions. However, the scalp and skull introduce resistance that can influence recorded signals. Therefore, the measured activity likely reflects cortical electrical field potentials rather than direct neural sources. This limits the precision with which we can interpret EEG data as a direct readout of cortical processes. Future work could improve spatial resolution by incorporating high-density EEG or invasive methods such as stereo-EEG (sEEG) or ECoG. Third, the reported correlations between beta-gamma PAC and motor impairment were based on cross-participant correlations. While informative at the group level, these do not capture individual variability over time. This is particularly important for establishing PAC as a real-time biomarker for applications such as adaptive DBS (aDBS), which requires sensitivity to within-subject fluctuations in symptom severity. Despite these limitations, our current results offer promising support for PAC as an objective biomarker of motor impairment in PD. Nevertheless, further work is needed to determine its reliability and specificity at the individual level. Future research should aim to include longer, real-world recordings and adopt within-subject analyses to more fully explore the potential of PAC as a dynamic and personalized biomarker for PD and its application in aDBS.

### Conclusions

This study identifies enhanced alpha/low beta-gamma PAC in both cortical and cortico-muscular networks as potential biomarkers for PD. Aberrant walking-related PAC interactions, particularly during levodopa withdrawal, suggest that increased beta-gamma coupling may contribute to motor and gait impairments in PD. The sensitivity of alpha/beta-gamma PAC to medication status highlights its potential utility for assessing therapeutic efficacy and informing targeted interventions. Furthermore, the application of non-invasive, dynamic monitoring, particularly during gait, highlights a promising avenue for the development of clinically relevant biomarkers. Such approaches may ultimately enable more precise, close loop systems for tracking and modulating neural and muscular activity in real time, with the goal of improving clinical outcomes in PD patients.

## Acknowledgements

We would like to present our acknowledgements to our patients for participating in this project, to Prof. Vladimir Litvak for his constructive suggestions on analysis.

## Data Availability

All relevant codes reported in the paper can be freely accessed without restriction (https://github.com/hyphenzhao/PD_PAC_Biomarkers). The raw data that support the findings of this study are available from the corresponding author upon reasonable request after approval of local ethics committee.

## Author Contributions

HZ: Writing – original draft, Visualization, Methodology, Investigation, Formal analysis, Data curation, Conceptualization, Funding acquisition. SHao: Writing – review & editing, Methodology, Visualization, Investigation, Data curation. PZ: Writing – review & editing, Visualization, Methodology, Data curation. SHe: Writing – review & editing, Methodology. LW: Writing – review & editing, Statistics. ZF: Writing – review & editing, Investigation, Data curation, Resources. LX: Investigation, Data curation, Resources. SZ: Data curation, Resources. WL: Data curation, Resources. XZ: Data curation, Resources. MW: Writing – review & editing. DL: Data curation, Resources. BS: Data curation, Resources. YL: Resources, Funding acquisition, Supervision. HT: Writing – review & editing, Methodology, Conceptualization, Supervision. CC: Writing – review & editing, Methodology, Conceptualization, Supervision, Resources, Funding acquisition.

