## Supplemental Materials for "Cortical and Cortico-Muscular Beta-Gamma Phase Amplitude Coupling during Different Locomotion Status and the Effects of Levodopa in Parkinson’s Disease"

### **Supplementary Materials**

*Recording Details*

All patients underwent scalp shaving as part of the surgery preparation procedure before EEG recordings. This step helped mitigate movement artifacts, as highlighted in Kline et al.'s study. Therefore, the EEG cap was assured to firmly attached on the scalp with additionally elastic net cap on top of EEG cap. This further assure the data quality by minimizing the movement artifact. Additionally, all cables were securely bounded to the participants' bodies to prevent cable movement artifacts. To ensure precise heel strike detection, we utilized two sets of equipment with different shoe sizes to accommodate participants' varying foot dimensions. This approach allowed us to select the shoe size that best fit each participant, ensuring that the heel position was optimally aligned with the embedded sensors. By fully covering the sensors, we minimized the risk of misalignment or incomplete sensor coverage, thereby enhancing the accuracy of heel strike detection. The system ensured mobility using a power supply through a battery-charged Power over Ethernet (PoE) switch.

*EEG and EMG analyses*

The primary focus of this study was on central lobe channels C1, C2, and Cz, which are associated with lower limb functionality and are also less impacted by movement and neck artifacts. This selection strategy was also supported by its use in lower limb-related tasks such as brain-computer interface (BCI) applications and motor imagery identifications


^1,2^. The previous studies demonstrated that these channels could mainly represent signals from M1


^3,4^. Data processing was conducted using the MNE library in Python


^5,6^. All signals were sampled at 512Hz. The data underwent 50Hz notch filtering and 1-250Hz band-pass filtering. The data was then average referenced, followed by independent component analysis (ICA) to remove non-brain artifacts. The 23 EEG channels were decomposed to 21 ICA components. ICA components were classified by ICLabel


^7^ method and only components marked as ‘brain’ would be included to minimize the noise effects. The number of removed components ranged from four to ten. Cleaned data were saved in two formats: raw filtered data for continuous analyses and step-segmented data based on detected heel-strikes from the of pressure sensors for stepping-related investigations. All statistical analyses were conducted using the statsmodels


^8^ and scipy


^9^ libraries in Python.

Step data segments with peak-to-peak amplitudes over 400 microvolts and deviating beyond four times the standard deviation from the mean were excluded to reduce noises and outliers. Visual inspection of the excluded epochs' ERP and TFR confirmed that the algorithms effectively removed those epochs containing large spikes and cross-spectrum synchronizations. A visual inspection of the simultaneously video recording corrected misclassified triggers, ensuring data accuracy. No obstruction was involved. The continuous walking data included turning for general walking analysis whilst heel strike segmented data excluded turning by visually inspection of the simultaneously video recording, i.e. only straight walking was analyzed in segmented data. Four participants cannot finish event one walking forth and back in OFF status, and one participant had FOG when ON medication finished walking session after taking levodopa. The FOG epochs in that session were excluded as those caused misclassification of steps. To ensure optimal data quality, inter-subject consistency, and computational efficiency, the first 20 clean steps from each side of each patient were selected for analysis. Time-frequency decomposition of step data was performed using a Morlet wavelet transform.

*Phase Amplitude Coupling*

In this study, Python based Tensorpac


^10^ was employed to investigate the interactions between neural oscillations at different frequencies. We specifically utilized Gaussian Copula PAC (GC-PAC)


^10^ due to its benefits in managing short-length data. The GC-PAC procedure is a semi-parametric method that estimates mutual information between the phase of one signal and the amplitude of another, providing a robust measure of cross-frequency coupling. Specifically, it involves transforming both phase (represented by sin(ϕ(t)) and cos(ϕ(t))) and amplitude (a(t)) signals into standard normal distributions using copula normalization. This step ensures that the estimation focuses solely on the dependence structure and is unaffected by the marginal distributions or amplitude scaling. Mutual information is then calculated using a parametric, bias-corrected Gaussian estimator:

$$gcPAC=I\left( a\left( t \right);\left[ \sin\left( \phi\left( t \right) \right),\cos\left( \phi\left( t \right) \right) \right] \right)\ldots(1)$$

GC-PAC has advantages over traditional PAC measures, as it avoids binning and is robust to differences in signal-to-noise (SNR) ratios and distributional shapes. This makes it particularly suitable for analyzing PAC between signals with different modalities (e.g., cortical EEG and muscular EMG) and across short time windows. By leveraging this methodology, PAC can reliably detect cortex-muscle interactions, minimizing biases introduced by noise or signal strength discrepancies. This robustness makes it well-suited for exploring phase-amplitude coupling in PD, a novel area of investigation that has yet to be extensively studied. GC-PAC is particularly advantageous for PAC estimation in segmented data, such as those related to individual walking steps. It is resistant to amplitude shifts, ensuring a stable and accurate representation of neural interactions. For step-related PAC, the data was segmented and then calculated across trials by:

$$r_{\mathrm{sx}}=c\left( \sin\left( \phi_{t} \right), a_{t} \right), r_{\mathrm{cx}}=c\left( \cos\left( \phi_{t} \right), a_{t} \right), and r_{\mathrm{sc}}=c\left( \sin\left( \phi_{t} \right),\cos\left( \phi_{t} \right) \right)\ldots\left( 2 \right)$$

$$\rho_{\mathrm{cl}}=\sqrt{\frac{r_{\mathrm{sx}}^{2}+r_{\mathrm{cx}}^{2}-2r_{\mathrm{sx}}r_{\mathrm{cx}}r_{\mathrm{sc}}}{1-r_{\mathrm{sc}}^{2}}}\ldots\left( 3 \right)$$

The circular-linear method offers superior temporal precision by analyzing PAC at each sampling point, unlike sliding window approaches that average over intervals and may obscure rapid fluctuations. This allows for the detection of transient changes and a detailed representation of PAC dynamics, capturing the nuanced temporal evolution of neural interactions


^11^. PAC was always calculated at the channel-pair level before averaging. Although Amplitude-to-Amplitude Coupling (AAC) results were not included in the main results due to lack of statistical significance, AAC was calculated using PyBispectrum


^12^. These findings, detailed in the appendix, aim to provide a comprehensive view and support future inquiries into AAC's potential roles in neural and muscular interactions (Supplementary Figure 3C).

The previous study by Yin et al.


^13^ identified beta-gamma PAC in the primary motor cortex as a critical biomarker for motor impairments in PD, specifically freezing of gait (FOG). The researchers found increased beta-broadband gamma PAC associated with FOG episodes and that DBS can reduce this coupling, alleviating FOG symptoms. Building on this, we began by using evidence-based analysis beta (12-30Hz) and broad gamma (40-200Hz) activity in EEGs. Based on the evidence-based result, the further data-driven approach was then conducted by using permutation. The permutation test of PAC initially identified a range of 7.5-11.5Hz and 93.0-117.0Hz significant area on cEEG (Supplementary Figure 3A). This provides initial clues for further phase identification when averaging alongside amplitude axis. Similar permutation was also conducted on cEEG-gEMG PAC. This specified a significant cluster of 13.0-14.5Hz and 135.0-200.0Hz (Supplementary Figure 3B). This suggested a higher amplitude range which was then used in averaging along amplitude axis.

*Data Segmentations*

In this study, two types of data were utilized for analysis: continuous time data and step-segmented data. Continuous time data were recorded over one minute while sitting, standing, or walking, respectively, capturing both EEG and EMG signals. Exactly one-minute continuous data was cropped from each locomotion status to control the data duration influence. PAC was firstly explored by this continuous time to compare the effect of levodopa and locomotion (applied to Figures 4A, 4B, 4C, 5A, 5B, 5C). Additionally, data were segmented based on heel strikes, as defined earlier. Notably, combining consecutive step-segmented data effectively transforms it into continuous time data with varying total lengths but consistent step numbers. During the permutation test, step data segments from different conditions were randomly swapped and combined to generate the null hypothesis distribution. Specifically, for frequency-level decomposition, the entire gait cycle (defined as the data between consecutive heel strikes) was used for the step-segmented permutation test (applied to Figures 4D, 4E, 5D, 5E, 5F, 5G).

The epoch data was interpolated in different processing steps depending on the analysis type to formalize the data length and offset velocity and frequency variations of walking steps. The basis linear interpolation is:

$$x_{i}=\frac{i}{N-1}\quad\left( i=0,1,\ldots,N-1 \right),x_{j}^{'}=\frac{j}{M-1}\quad\left( j=0,1,\ldots,M-1 \right)\ldots\left( 4 \right)$$

$$\tilde{d}\left( x_{j}^{'} \right)=d\left( x_{i} \right)+\frac{x_{j}^{'}-x_{i}}{x_{i+1}-x_{i}}\left( d\left( x_{i+1} \right)-d\left( x_{i} \right) \right),\quad\text{for }x_{i}\leq x_{j}^{'}\leq x_{i+1}\ldots\left( 5 \right)$$

where x_i_ represents the original time index and x_j_ denotes the new time index. The new time index was standardized to 2048 samples to represent a 2-second gait cycle, and the final time indices were expressed as percentages of the gait cycle. In power analysis, the TFR was firstly decomposed on original data:

$$\text{TFR}\left( t,f \right)=\left| \int_{-\infty}^{\infty} x\left( \tau\right) \psi_{f}^{*}\left( \tau-t \right) d\tau\right|^{2},\psi_{f}\left( t \right)=A e^{-\frac{t^{2}}{2\sigma_{t}^{2}}} e^{2\pi ift}\ldots\left( 6 \right)$$

And each frequency decomposition was treated as a one-dimensional array to interpolated to 2 seconds (representing a full gait cycle of 100%). For event-related PAC, the original signals were transferred into several signal component by phase filters and amplitude filters. Let x(t) be the original signal, for each frequency band i with lower cutoff $f_{\text{low}}^{\left( i \right)}$and upper cutoff$f_{\text{high}}^{\left( i \right)}$, define the filter transfer function $H_{i}\left( \omega\right)$. The filtered signal is given by:

$$x_{i}\left( t \right)=\mathcal{F}^{-1}\left\{ H_{i}\left( \omega\right)\mathcal{F}\{x\left( t \right)\} \right\},x\left( t \right)\quad\to\quad\{x_{i}\left( t \right){\}}_{i=1}^{N}\ldots\left( 7 \right)$$

Each filtered signal was interpolated to a fixed duration of 2 seconds (representing a full gait cycle of 100%), after which the sine and cosine components were computed and their correlation calculated.

*References*

### 1. Gordleeva, S. Y. *et al.* Real-Time EEG–EMG Human–Machine Interface-Based Control System for a Lower-Limb Exoskeleton. *IEEE Access* **8**, 84070–84081 (2020).

### 2. Ferrero, L. *et al.* Brain Symmetry Analysis during the Use of a BCI Based on Motor Imagery for the Control of a Lower-Limb Exoskeleton. *Symmetry* **13**, 1746 (2021).

### 3. Liu, S. *et al.* Interference of unilateral lower limb amputation on motor imagery rhythm and remodeling of sensorimotor areas. *Frontiers in Human Neuroscience* **16**, (2022).

### 4. AL-Quraishi, M. S. *et al.* Bimodal Data Fusion of Simultaneous Measurements of EEG and fNIRS during Lower Limb Movements. *Brain Sciences* **11**, 713 (2021).

### 5. Gramfort, A. MEG and EEG data analysis with MNE-Python. *Frontiers in Neuroscience* **7**, (2013).

### 6. Larson, E. *et al.* MNE-Python. (2024).doi:10.5281/ZENODO.592483

### 7. Pion-Tonachini, L., Kreutz-Delgado, K. & Makeig, S. ICLabel: An automated electroencephalographic independent component classifier, dataset, and website. *NeuroImage* **198**, 181–197 (2019).

### 8. Seabold, S. & Perktold, J. statsmodels: Econometric and statistical modeling with python. *9th Python in Science Conference* (2010).

### 9. Virtanen, P. *et al.* SciPy 1.0: fundamental algorithms for scientific computing in Python. *Nature Methods* **17**, 261–272 (2020).

### 10. Combrisson, E. *et al.* Tensorpac: An open-source Python toolbox for tensor-based phase-amplitude coupling measurement in electrophysiological brain signals. *PLOS Computational Biology* **16**, e1008302 (2020).

### 11. Voytek, B., D’Esposito, M., Crone, N. & Knight, R. T. A method for event-related phase/amplitude coupling. *Neuroimage* **64**, 416–424 (2013).

### 12. Binns, T. S., Pellegrini, F., Jurhar, T. & Haufe, S. PyBispectra v1.1.0. (2023).doi:10.5281/ZENODO.8377820

### 13. Yin, Z. *et al.* Cortical phase-amplitude coupling is key to the occurrence and treatment of freezing of gait. *Brain* **145**, 2407–2421 (2022).

### **Supplementary Figure**

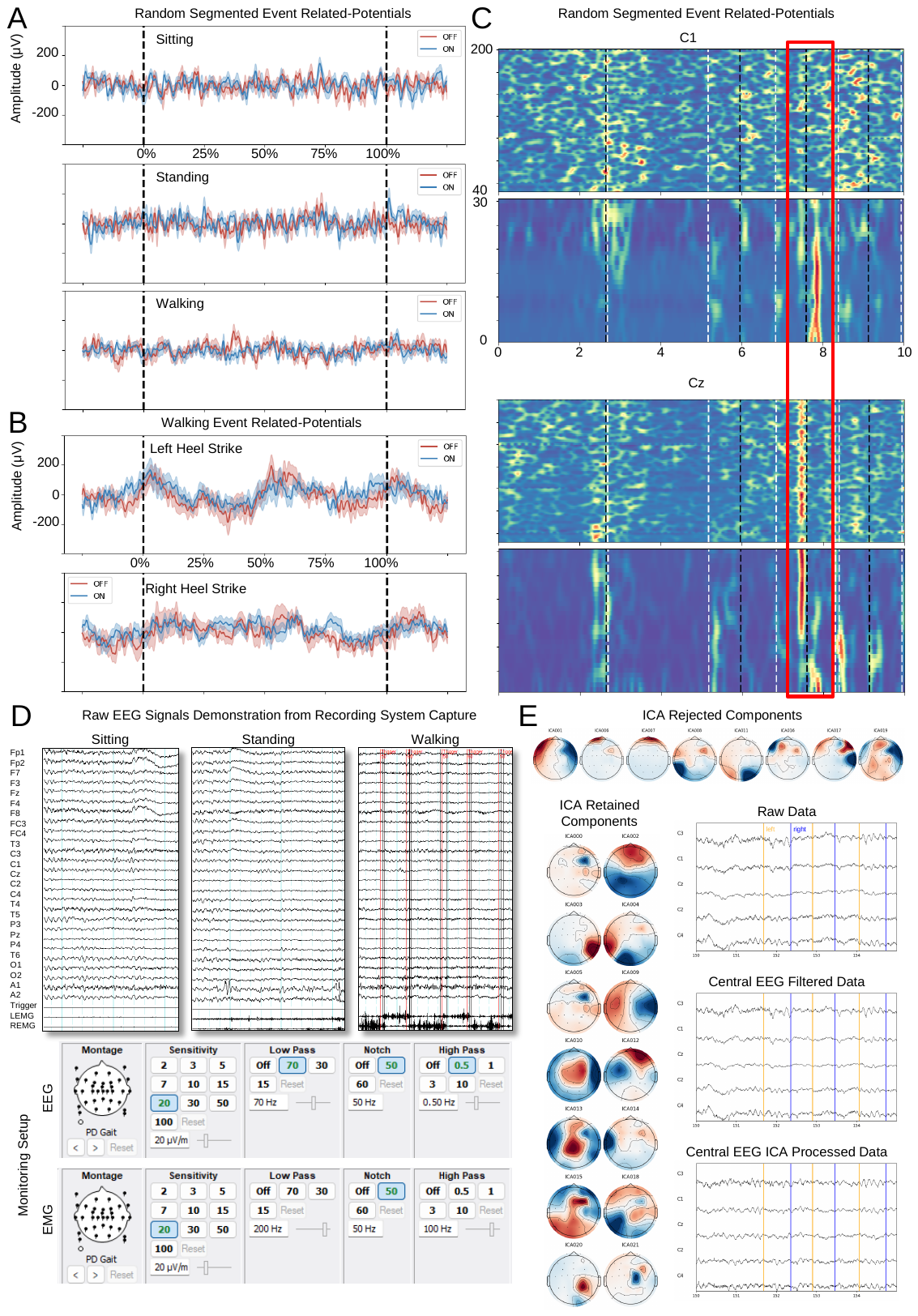

**Supplementary Figure 1** Event-Related Potentials (ERP) for sitting, standing, and walking demonstrates the data quality of recordings. (A) Continuous central EEG data from sitting, standing, and walking were randomly segmented into epochs to simulate ERP responses. The resulting confidence intervals indicated acceptable variability. (B) ERPs from real walking epochs, segmented using foot-triggered events, exhibited slightly larger confidence intervals; however, the signal-to-noise ratio remained within a reasonable range. (C) A demonstration of TFR on single 10 seconds decomposition shows that the broadband activation can be easily found. These types of noises were ensured excluded from analysis by visually inspection. (D) Raw EEG signals captured using the Profusion EEG 6 system demonstrated reliable recordings across sitting, standing, and walking conditions, particularly in the central lobes. The vertical lines in the walking recordings represent triggers from the real-time pressure monitoring system, confirming precise alignment between these triggers and the corresponding EMG activity. (E) Example of ICA processing illustrating component rejection and retention. The figure displays the raw signal, the filtered signal, and the signal after ICA processing.

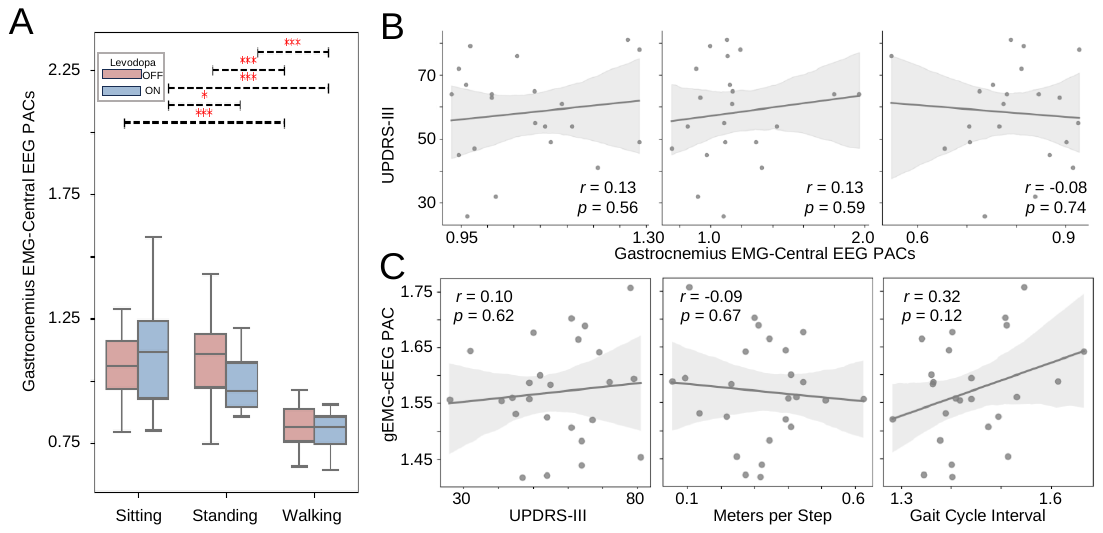

**Supplementary Figure 2** The figure shows reverse beta-gamma PAC between cEEG and gEMG (gEMG-cEEG). The reverse beta-gamma PAC (gEMG-cEEG) indicates that clinically relevant PAC is evident only when directed from cEEG to gEMG. (A) Under sitting, standing, and walking conditions, gEMG-cEEG PAC showed no significant differences between levodopa states and exhibited a pattern distinct from cEEG-gEMG PAC. (B) Beta-gamma gEMG-cEEG PAC did not correlate with MDS-UPDRS-III scores in any locomotion state. (C) Permutation tests on walking epochs of beta-gamma gEMG-cEEG PAC failed to identify any significant frequency differences. These results further support that the direction of PAC—from cEEG to gEMG—is critical, effectively ruling out the influence of EMG and movement noise in EEG recordings.

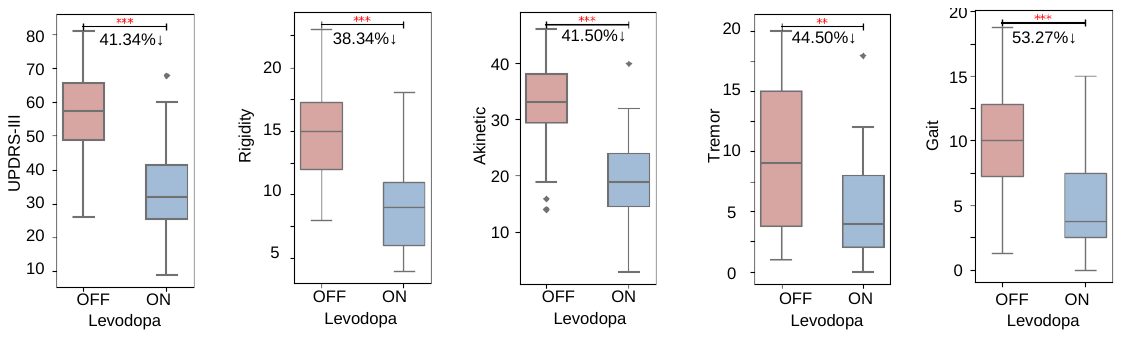

**Supplementary Figure 3** This plot compares the MDS-UPDRS-III total score, along with the Rigidity, Akinesia, Tremor, and Gait subsections, between OFF and ON states. The percentage improvement in scores from OFF to ON state is shown, with statistical significance indicated for each measure.

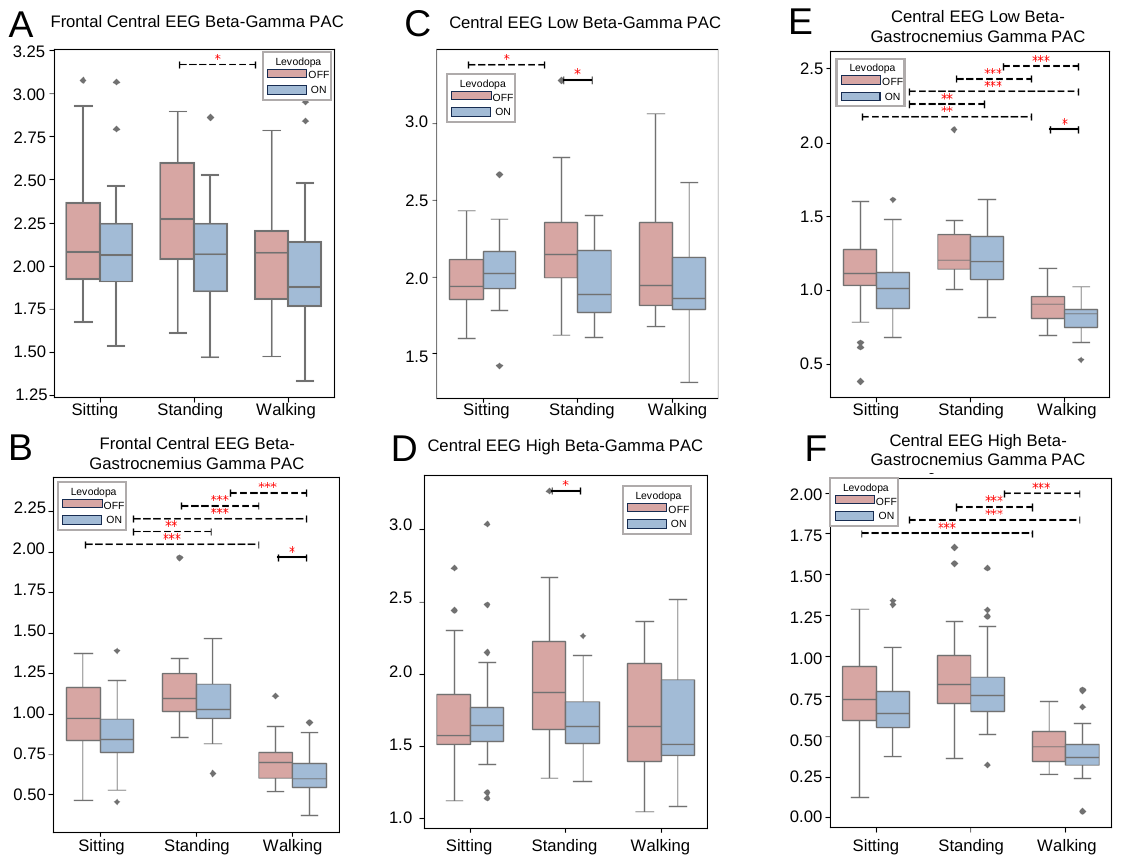

**Supplementary Figure 4** Phase-Amplitude Coupling (PAC) analysis of Frontal and Central EEG with Gastrocnemius EMG across various frequency bands. (A) Frontal Central EEG Beta-Gamma PAC during different locomotion and medication statuses. (B) Frontal Central EEG Beta-Gastrocnemius EMG Gamma PAC across locomotion and medication conditions. (C) Central EEG Low Beta-Gamma PAC analysis across different conditions. (D) Central EEG High Beta-Gastrocnemius EMG Gamma PAC across locomotion and medication statuses. (E) Central EEG Low Beta-Gastrocnemius EMG Gamma PAC analysis across locomotion and medication conditions. (F) Central EEG High Beta-Gastrocnemius EMG Gamma PAC analysis during different locomotion states and medication conditions. The Frontal Central EEG did not show significant sensitivity to medication or gesture, in contrast to Central EEG, which exhibited greater responsiveness. Additionally, the Low and High Beta bands of Central EEG did not demonstrate distinct, independent patterns from the Broad Beta band. Consequently, this study primarily focuses on Beta-Gamma PAC in Central EEG, with further data-driven methods applied to explore the optimal frequency bandwidth.

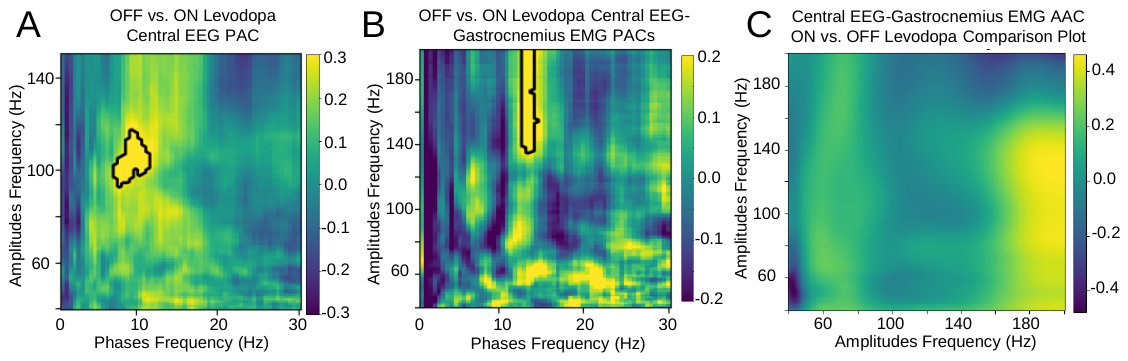

**Supplementary Figure 5** PAC and Amplitude-to-Amplitude Coupling (AAC) decompositions: (A) Permutation test on frequency decomposed walking cortical PAC identifies significantly different areas between OFF and ON levodopa states, specifically around phase frequencies of 7.5-11.5Hz and amplitude frequencies of 93.0-117.0Hz. (B) Permutation testing identifies significant walking cortico-muscular PAC differences between 13.0-14.5Hz (phase) and 135.0-200.0Hz (amplitude) frequencies, with higher values observed in the OFF levodopa state. Significant areas are highlighted by black contours. (C)This figure illustrates the AAC between EEG and EMG signals within the high-frequency range of 40-200 Hz from the first 20 steps during walking of ON vs. OFF levodopa status. The analysis shows regions of high coupling, primarily around 40-160 Hz for EEG and 160-180 Hz for EMG. However, these couplings did not reach statistical significance.

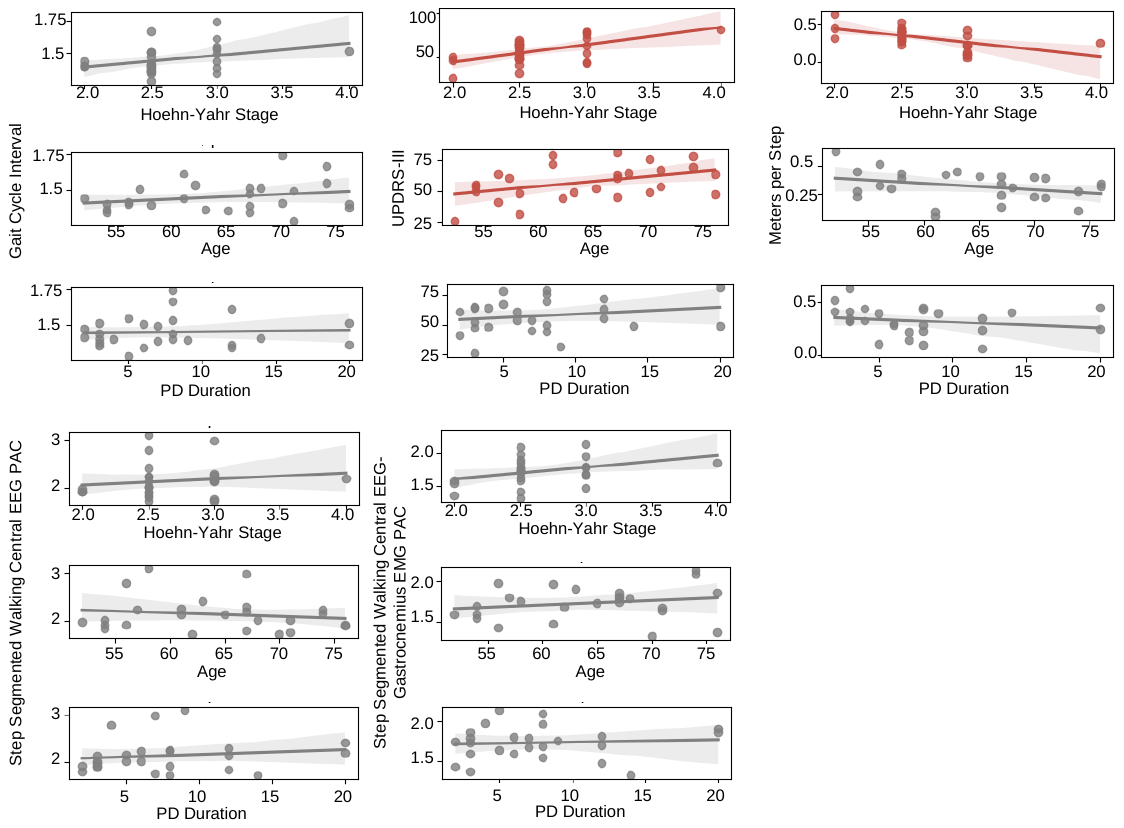

**Supplementary Figure 6** We conducted additional analyses to examine the correlations of Hoehn-Yahr (HY) stage, age, and PD duration with Gait Cycle Interval, MDS-UPDRS-III, Meters per Step, Step-Segmented Walking Cortical PAC, and Step-Segmented Walking Cortico-Muscular PAC. Among these, only HY stage showed significant correlations with MDS-UPDRS-III and Meters per Step, while age was also correlated with MDS-UPDRS-III, as expected. No significant correlations were observed for other parameters. All p-values were corrected for false discovery rate (FDR). These findings suggest that changes in PAC were not influenced by age, PD duration, or HY stage.
